# Using DEPendency of association on the number of Top Hits (DEPTH) as a complementary tool to identify novel risk loci in colorectal cancer

**DOI:** 10.1101/2022.11.24.22282734

**Authors:** John Lai, Chi Wong, Daniel F. Schmidt, Miroslaw Kapuscinski, Karen Alpen, Robert J. MacInnis, Daniel D. Buchanan, Aung K. Win, Jane Figueiredo, Andrew T. Chan, Tabitha A. Harrison, Michael Hoffmeister, Emily White, Loic Le Marchand, Ulrike Peters, John L. Hopper, Enes Makalic, Mark A. Jenkins

## Abstract

**Background:** DEPendency of association on the number of Top Hits (DEPTH) is an approach to identify candidate risk regions by considering the risk signals from over-lapping groups of sequential variants across the genome.

**Methods:** We conducted a DEPTH analysis using a sliding window of 200 SNPs to colorectal cancer (CRC) data from the Colon Cancer Family Registry (CCFR) (5,735 cases and 3,688 controls), and GECCO (8,865 cases and 10,285 controls) studies. A DEPTH score >1 was used to identify risk regions common to both studies. We compared DEPTH results against those from conventional GWAS analyses of these two studies as well as against 132 published risk regions.

**Results:** Initial DEPTH analysis revealed 2,622 (CCFR) and 3,686 (GECCO) risk regions, of which 569 were common to both studies. Bootstrapping revealed 40 and 49 likely risk regions in the CCFR and GECCO data sets, respectively. Notably, DEPTH identified at least 82 likely risk regions that would not be detected using conventional GWAS methods, nor had they been identified in previous CRC GWASs. We found four reproducible risk regions (2q22.2, 2q33.1, 6p21.32, 13q14.3), with the HLA locus at 6p21 having the highest DEPTH score. The strongest associated SNPs were rs762216297, rs149490268, rs114741460, and rs199707618 for the CCFR data, and rs9270761 for the GECCO data.

**Conclusion:** DEPTH can identify novel likely risk regions for CRC not identified using conventional analyses of much larger datasets.

**Impact:** DEPTH has potential as a powerful complementary tool to conventional GWAS analyses for identifying risk regions within the genome.

## Introduction

Colorectal cancer (CRC) is the third most diagnosed cancer in the Western World in both men and women, as well as accounting for the third highest cancer-related deaths (1,2). Notably, CRC mortality rates have been decreasing for the past few decades (1,2) driven in part by higher uptake of screening and the removal of polyps (3-6). Consequently, many national guidelines recommend that general CRC screening start from 45-50 years of age to reduce CRC mortalities as early intervention is an effective tool. Identifying individuals with a high risk of developing CRC may further improve CRC mortality as higher risk individuals may benefit from having earlier or more frequent CRC screening.

One CRC risk factor that has been of great interest is a genetic predisposition to develop this disease. There is a strong genetic component underlying CRC risk, with an estimate that the heritability of CRC is 28% for men, and 45% for women (7). Indeed, genome wide association studies (GWASs) have now identified 132 loci that are associated with CRC risk in individuals of European ancestry (8-12). Conventional GWASs typically assess the correlation of a single nucleotide polymorphism (SNP) with a phenotype independently, and then correct for multiple testing to identify SNPs that are significantly associated with the phenotype. However, this approach cannot account for SNPs that have a correlated association which may bias the results given the correction is overly conservative for SNPs that have a correlated association. As such, we have recently developed a method termed DEPendency of association on the number of Top Hits (DEPTH) to identify SNPs that are associated with a phenotype that assesses risk using multiple SNPs at the locus, rather than assessing SNP risk individually (13). Notably, we have applied DEPTH analyses to a prostate cancer GWAS data set and identified 112 novel prostate cancer risk regions (14).

Here, we have used DEPTH to identify novel CRC risk regions that are not as readily detected by conventional GWAS approaches. We compare our DEPTH results against conventional GWAS analyses on the same data sets, as well as comparing our DEPTH results against known CRC risk loci. We report in this study that DEPTH can identify novel high-confidence risk loci, and that this method is a useful complementary discovery tool for potential risk regions.

## Materials and Methods

### Study subjects

DEPTH analyses were carried out on two independent CRC data sets—the Colon Cancer Family Registry (CCFR) data set, and the Genetics and Epidemiology of CRC Consortium (GECCO) data set. The initial CCFR data set comprises 11,489 participants (7,151 cases and 4,338 controls) from five separate GWAS data sets, and the initial GECCO data set comprises 20,320 participants (9,498 cases and 10,822 controls) (Supplementary Table 1). Further information about CCFR and GECCO participants can be found in previous publications (11,12,15). Participants provided written informed consent by their respective Institutional Review Boards (need info), and self-reported race as being White.

### Genotyping quality control (QC) and data harmonisation

The initial imputed genotype data sets used in this study comprised of between 39,127,678 to 39,222,163 SNPs (Supplementary Table 2). QC and imputation had already been carried out by the respective consortia for these data sets (11,12,15,16). However, further QC was applied to ensure there is a baseline level of consistency between the data sets (data harmonisation) – see Supplementary Methods. QC and data harmonisation were primarily carried out using PLINK v1.9 (17) and custom Perl scripts (Supplementary Methods and Supplementary Figure 1). All chromosomal positions/regions specified in this study refer to genome build GRCh37/hg19.

### DEPTH SNPs and participants

Typical GWAS QC was applied to the CCFR and GECCO datasets, plus an extra QC step that removed SNPs that had an odds ratio or *P* value that differed by more than 20% when comparing logistic regression results that adjusted for sex and the first four principal components, against logistic regression results that adjusted for sex but not the first four principal components (Supplementary Figure 3). This extra QC step was performed to remove SNPs that may be affected by principal components, as DEPTH analyses cannot adjust for principal components. The number of SNPs and patients that were removed after each step are detailed in Supplementary Tables 2 and 4. In total 7,234,408 SNPs on 10,846 CCFR participants (5,735 cases and 3,688 controls), and 19,150 GECCO participants (8,865 cases and 10,285 controls) were retained for DEPTH analyses following genotyping QC and data harmonisation.

### DEPTH analyses

All DEPTH analyses were carried out using the University of Melbourne’s high-performance computing system (18). Risk regions were defined as having a posterior log-odds in favour of association above 1.0. This risk value was based on a previous DEPTH study which suggests that the maximum 95^th^ percentile of the null distribution equals approximately 1.0 across the genome (14). DEPTH analyses were carried out in three stages, whereby the aim of the first stage was to identify potential common regions of CRC risk between the CCFR and GECCO data sets. – see Supplementary Figure 2 for a description of common regions. The second stage then evaluated candidate higher-confidence regions (95% confidence interval) using 100 simulations on the common risk regions that were identified in the first stage. The third stage entailed carrying out a more rigorous DEPTH simulation analysis using 1,000 bootstrap iterations on risk regions identified in stage 2 that was supported at the 95% confidence level. DEPTH was carried out using a sliding window of 200 SNPs for all stages. Genes that are located within DEPTH risk regions were identified by scanning DEPTH regions against the UCSC Genes table (GRCh37/hg19) (19) using a custom Perl script.

### Comparison of DEPTH results with conventional GWAS

Conventional GWAS logistic regression analysis of both the CCFR and GECCO data sets were carried out on 14,285,390 SNPs that were common to both data sets. Apart from the last QC step, these SNPs underwent the same QC and data harmonisation as the DEPTH data sets (data sets from Step 12 of Supplementary Table 2). Logistic regression was carried out using PLINK v1.9 (17), adjusting for sex and the first four principal components. A *P* value less than 5 × 10^−8^ was used to define a significantly associated SNP. GWAS summary statistics were uploaded to the Functional Mapping and Annotation of Genome-Wide Association Studies (FUMAR) platform (20) to identify and annotate SNPs that are associated with CRC risk.

The 50 CCFR DEPTH risk regions and the 66 DEPTH risk regions in the GECCO data sets that were identified at the 95^th^ percentile using 100 simulations were then compared with conventional GWAS analysis results using a custom Perl script. A comparison of the 40 CCFR and 49 GECCO DEPTH risk regions at the 95^th^ percentile using 1,000 simulations with conventional GWAS results was also carried out.

A comparison of DEPTH risk regions against previously published risk loci from conventional CRC GWASs was also carried out. This was performed as above but using the 132 risk loci (Supplementary Table 3) from the Law et. al. and Huyghe et. al. studies (10,11). However, of the 132 known CRC risk SNPs, only 98 risk SNPs are found in our DEPTH data sets comprising 7,234,408 SNPs and thus, only these 98 SNPs were assessed when comparing logistic regression risk SNPs against DEPTH peaks and the 98 known CRC risk SNPs (Supplementary Table 3). It was not possible to use LD SNPs for the remaining 34 known CRC risk SNPs because LD SNPs were removed during the QC process to facilitate feasible computational time for the bootstrap iteration step.

### Ranking SNPs within the HLA risk locus using penalised regression

The HLA region was chosen for further analysis given it was the most reproducible common loci detected between the CCFR and GECCO datasets after Stage 3 analysis. In total, 381 SNPs that are located within the HLA risk locus were used for lasso logistic regression and random forest analyses in both the CCFR and GECCO data sets. The random forest supervised machine learning algorithm provides an automatic relevance ranking of all SNPs that were used to fit the model. With lasso logistic regression, the order in which the SNPs entered the regression model was taken as a surrogate for their rank. For example, the first SNP that entered the model was ranked top, the second SNP that entered the regression model was ranked second, etc.

## Results

### Identification of CRC risk regions by DEPTH analyses

The first stage of DEPTH analysis identified 2,622 risk regions in the CCFR data set, 3,686 risk regions in the GECCO data set (Table 1 and Supplementary Table 5), and 569 risk regions that were common to both the CCFR and GECCO data sets (Supplementary Table 6). The second stage of DEPTH analysis indicated that of these 569 common risk regions, 50 (8.8%) and 66 (11.6%) remained significantly associated with risk after 100 bootstrap iterations at the 95% confidence level in the CCFR and GECCO data sets, respectively, with 5 regions that were common to both data sets (Table 1 and Supplementary Table 7). The third stage of DEPTH analysis indicated that of the 50 CCFR and 66 GECCO risk regions from stage 2, 40 (80.0%) and 49 (76.6%) remained significantly associated with risk after 1,000 bootstrap iterations at the 95% confidence level in the CCFR and GECCO data sets, respectively, with four regions (chr2:143,560,338 – 143,668,969 (2q22.2), chr2:201,851,477 – 201,991,983 (2q33.1), chr6:32,421,184 – 32,682,590 (6p21.32), and chr13:52,434,734 – 52,788,187 (13q14.3)) that were common to both data sets (Table 1 and Supplementary Table 8). Notably, three ((chr2:143,560,338 – 143,668,969 (2q22.2), chr2:201,851,477 – 201,991,983 (2q33.1), and chr13:52,434,734 – 52,788,187 (13q14.3)) of the four regions were not discovered by previous GWAS studies, and none were detected using logistic regression analysis using the same datasets (Supplementary Table 8).

### DEPTH analyses identify risk regions that are not detected by conventional GWAS

Conventional GWAS using logistic regression analyses revealed four (CCFR) and nine (GECCO) SNPs that are associated with CRC risk (Supplementary Table 9 and Supplementary Figure 4), and FUMAR analysis indicates that these SNPs map to six independent risk loci (CCFR: rs13200613 at 6q27, rs2391397 at 7p15.2, rs4325396 at 13q11, rs4553118 at 1p36.33; GECCO: rs6983267 at 8q24.21, rs79207432 at 15q13.3) (Table 2). A comparison of these six lead conventional GWAS logistic regression risk SNPs from our DEPTH data sets against 98 of the 132 known CRC risk loci in our DEPTH dataset indicates that only two (rs6983267 and rs79207432) of the 98 loci (1.52%) could be detected in our CCFR and GECCO data sets (Table 2 and Supplementary Table 3).

Similar comparative analysis of our CCFR and GECCO conventional GWAS results against DEPTH first stage results indicates that only one (rs13200613) of the six lead logistic regression risk SNPs are located within the 569 common DEPTH risk regions (Supplementary Table 6). Notably, these three risk loci were also detected in DEPTH stage 2 and 3 results (Supplementary Tables 7 and 8). Thus, most DEPTH risk loci (568 of 569 in the first stage, 110 of 111 in the second stage, and 84 of 85 in the third stage) cannot be detected by conventional GWAS analyses in our CCFR and GECCO data sets (Figure 1 and Supplementary Tables 6-10).

**Figure 1.**
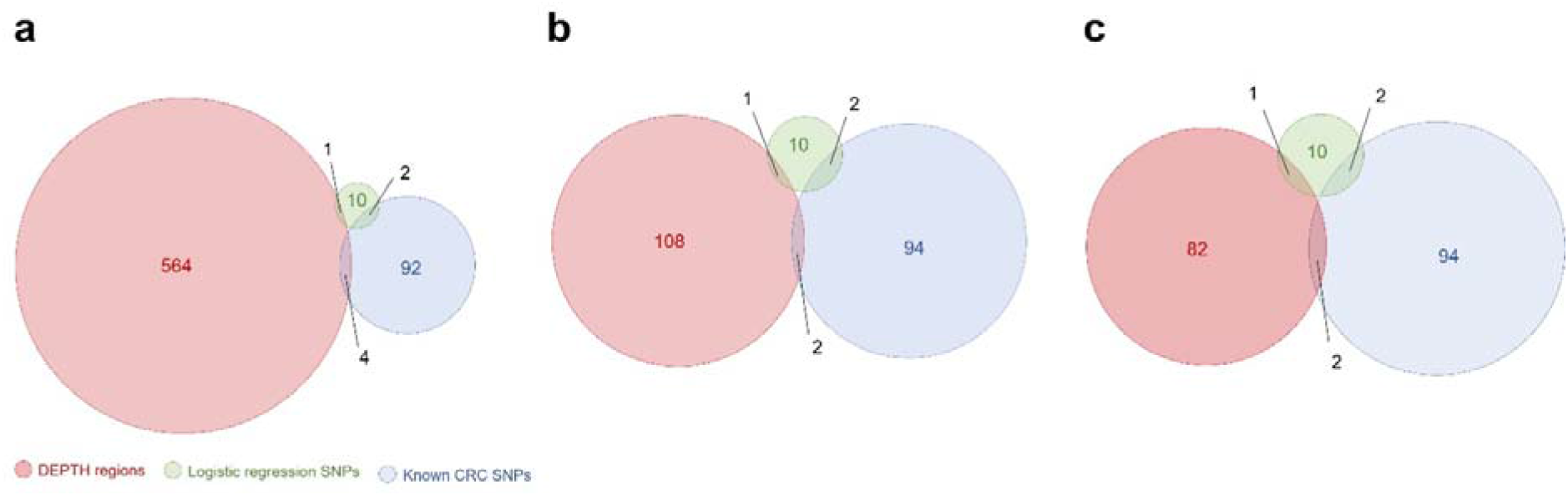
Venn diagram showing how many of the DEPTH risk regions (red circles) were detected by conventional GWAS analyses using the same data set (green circles) or were found in previous CRC GWASs (blue circles). Analysis was compared across results from the first ***(a)***, second ***(b)***, and third ***(c)*** stages of DEPTH analyses.

A comparative analysis was also carried out to assess how many of the 98 known CRC risk loci that are in our DEPTH data set are also detected by DEPTH analysis. Our results indicate that four (4.1%), two (2.0%), and two (2.0%) of the 98 known risk loci was detected by the first, second, and third stages of DEPTH analysis, respectively (Figure 1 and Supplementary Table 3). Thus, most of the DEPTH risk regions have not been detected by previous studies that use conventional GWAS approaches. Collectively, DEPTH identified 564 (first stage), 108 (second stage) and 82 (third stage) novel risk regions that could not be detected using conventional GWAS approaches in our CCFR and GECCO data sets, nor were they identified in previous CRC GWASs (Figure 1).

### Analysis of the Human Leukocyte Antigen (HLA) locus

The HLA risk locus spanning chr6:32,421,184-32,682,590 at 6p21 (Figure 1) was found to be most significantly associated with CRC risk at all stages of DEPTH analyses and was consistently significant in both CCFR and GECCO datasets (Table 3). As such, this region was selected for penalised regression analysis to determine the main SNPs that may confer risk in this region. Our results indicate that the most associated SNPs that were identified by all three methods were rs762216297, rs149490268, rs114741460, and rs199707618 for the CCFR data set, and rs9270761 for the GECCO data set, all of which cover the same HLA region (Figure 2 and Supplementary Table 10).

**Figure 2.**
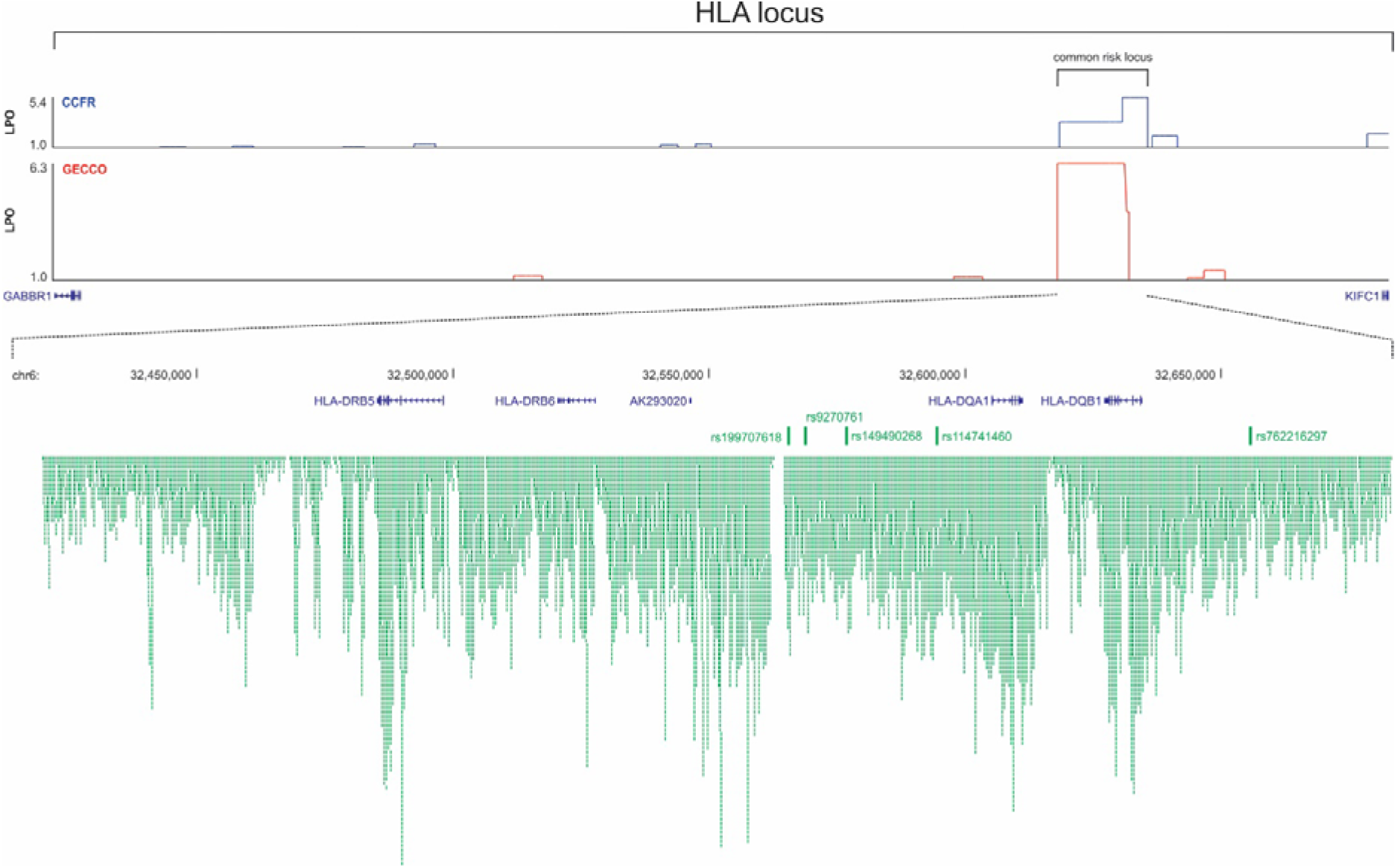
Diagram of the HLA locus (bordered by the *GABBR1* and *KIFC1* genes) showing a common DEPTH risk region. Loci with a log-posterior odds (LPO) above one are classified as risk regions. Also shown is a detailed view of the common DEPTH risk region that encompasses the *HLA-DRB5, HLA-DRB6, AK293020, HLA-DQA1*, and *HLA-DQB1* genes, as well as the five main SNPs (rs199707618, rs9270761, rs149490268, rs114741460, and rs762216297) driving risk in this region. Chromosomal positions above the genes refer to GRCh37/hg19 genome build. The density of common SNPs in the common risk region is shown by the green blocks.

## Discussion

Since the first GWAS was proposed in 1996 (21), this method has been used extensively to identify low penetrant but common SNPs that are associated with a wide range of complex phenotypes. For CRC, 132 SNPs have been found to be associated with the risk of developing this disease in individuals of European ancestry (8,10-12), of which 98 SNPs could be tested in this study. Novel SNPs that are associated with CRC risk are increasingly more difficult to identify using conventional GWAS approaches. As such, conventional GWASs have adopted various techniques such as increasing sample sizes, meta-analysis, and using imputed SNPs, to identify novel lower penetrant SNPs in subsequent GWASs. Nevertheless, these approaches still assess for association by using one SNP at a time. However, this approach is overly conservative because it uses excessively stringent Bonferroni thresholds because they assume statistical independence of SNPs, even though there is linkage disequilibrium. In this study, we have applied a novel alternative approach called (DEPTH), whereby we have investigated CRC risk regions by using a sliding window of 200 SNPs to collectively identify risk regions in two CRC GWAS data sets (CCFR and GECCO). DEPTH is a hypothesis generation algorithm based on a novel feature ranking algorithm incorporating data re-sampling and stability selection (13). The utility of DEPTH in discovering novel risk regions using GWAS data sets is highlighted by our identification of 112 novel prostate cancer risk regions, as well as five known risk regions that have only been identified by much larger conventional GWAS analyses (14). Had we limited our analysis to the conventional GWAS method applied to both data sets, we would have only found 6 independent loci.

Here, we performed DEPTH analysis in three stages due to the computational demands of DEPTH. This approach ultimately led to the identification of 37 (CCFR) and 48 (GECCO) risk regions that were not detected by conventional GWAS analyses, nor were they discovered in previous CRC GWASs, demonstrating the novelty of our approach.

Interestingly, the most highly associated DEPTH risk region (6p21.32 at chr6:32,421,184-32,682,590) was located within the human leukocyte antigen (HLA) locus, which is defined as being between the *GABBR1* and *KIFC1* genes (22). Notably, this CRC risk region could not be detected until only recently when Huyghe et al and Law et. al. carried out a meta-analysis that includes new data sets that had not been used in previous GWASs (10,11). However, we were able to identify the HLA risk loci despite only having 5,735 cases and 3,688 controls for the CCFR data set, and 8,865 cases and 10,285 controls for the GECCO data set, whereas Law et al. had 34,627 cases and 71,379 controls. Thus, DEPTH has a sensitivity that is much greater than conventional GWAS approaches for the same sample size. Further, the SNP (rs9271770) at the HLA risk region in Law et. al. is located directly within our DEPTH 6p21.32 risk region, even though this SNP is not in our data set. Thus, we were able to identify this risk loci without using rs9271770. The other SNP (rs3131043) at the HLA risk region in the Law et. al. study was not located within any of our DEPTH risk regions. Interestingly, the main SNPs driving risk at this locus are different between the two data sets (rs762216297, rs149490268, rs114741460, rs199707618 for CCFR, and rs9270761 for GECCO). The correlation (r^2^) and linkage disequilibrium (D prime) between rs199707618 and rs9270761 is 0.02 and 1—the discrepancy possibly due to rs9270761 having a very low minor allele frequency. This finding further highlights the utility of DEPTH in that it is not solely reliant on one SNP as it might be due to non-linear associations and epistasis.

The HLA DEPTH risk region in this study harbours five genes (*HLA-DRB5, HLA-DRB6, AK293020, HLA-DQA1, HLA-DQB1*), whereas the HLA locus collectively comprises over 130 protein coding genes that are important in regulating the immune system (22). While the HLA locus and the immune system has been known to be important in a wide range of diseases such as diabetes, rheumatoid arthritis and various autoimmune disorders (22), only recently has the role of the immune system been intensively studied for its role in cancer, and the potential manipulation of this system in treating cancer through immunotherapy (23). It is tempting to speculate whether the HLA DEPTH risk region and the five genes located within would be of value in pharmacogenomics, or as a therapeutic target.

In conclusion, we show using two CRC GWAS data sets (CCFR and GECCO) that more risk regions can be identified using an alternative approach (DEPTH) without needing to increase sample sizes. In fact, this study reduced the number of participants compared to the original CCFR and GECCO data sets due to an extra layer of QC and data harmonisation that was applied to this study. Finally, we designed DEPTH as an exploratory tool to identify novel risk loci, with the expectation that other methods can later be used to validate these loci. For example, in this study we used penalised regression to identify the main SNPs driving risk at the HLA locus, although other methods such as bioinformatic and/or functional analyses of the SNPs at these risk loci may also inform on the biological reasons driving the risk association. We expect DEPTH will provide great utility to the GWAS field as a complementary tool to identify novel loci that are associated with complex phenotypes using existing GWAS data sets.

## Data Availability

All data produced are available online at the NIH NLM dbGap repository.

## Figure Legends

**Supplementary Figure 1.**
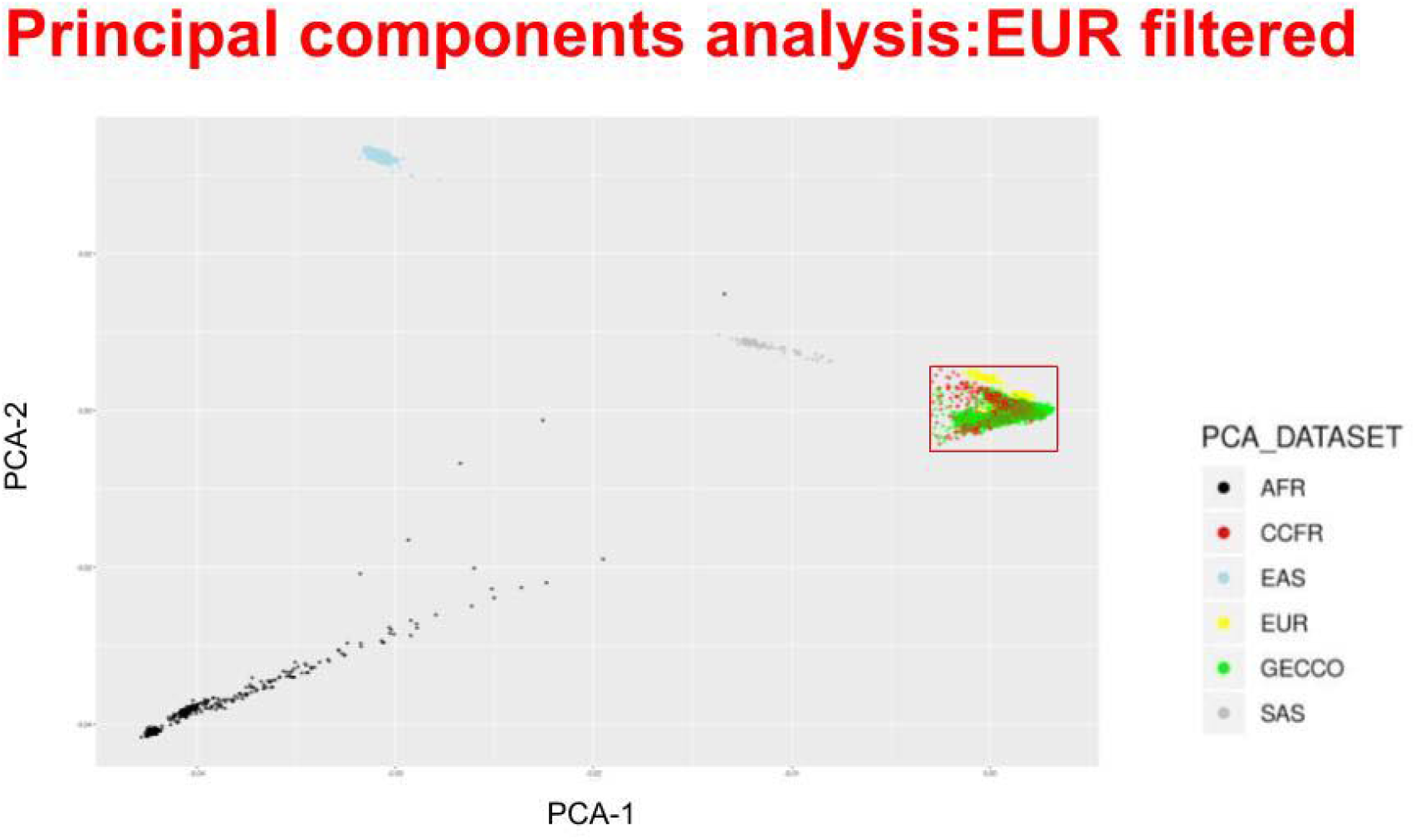
Filtering European patients based on the first two principal components (PCA-1 and PCA-2). Ethnic clusters were defined using the 1000 Genomes Project data set of African (AFR), East Asian (EAS), European (EUR), and South-East Asian (SAS) samples. CCFR (red dots) and GECCO (green dots) patients were only selected for further analyses if they clustered with the European population (within the red box).

**Supplementary Figure 2.**
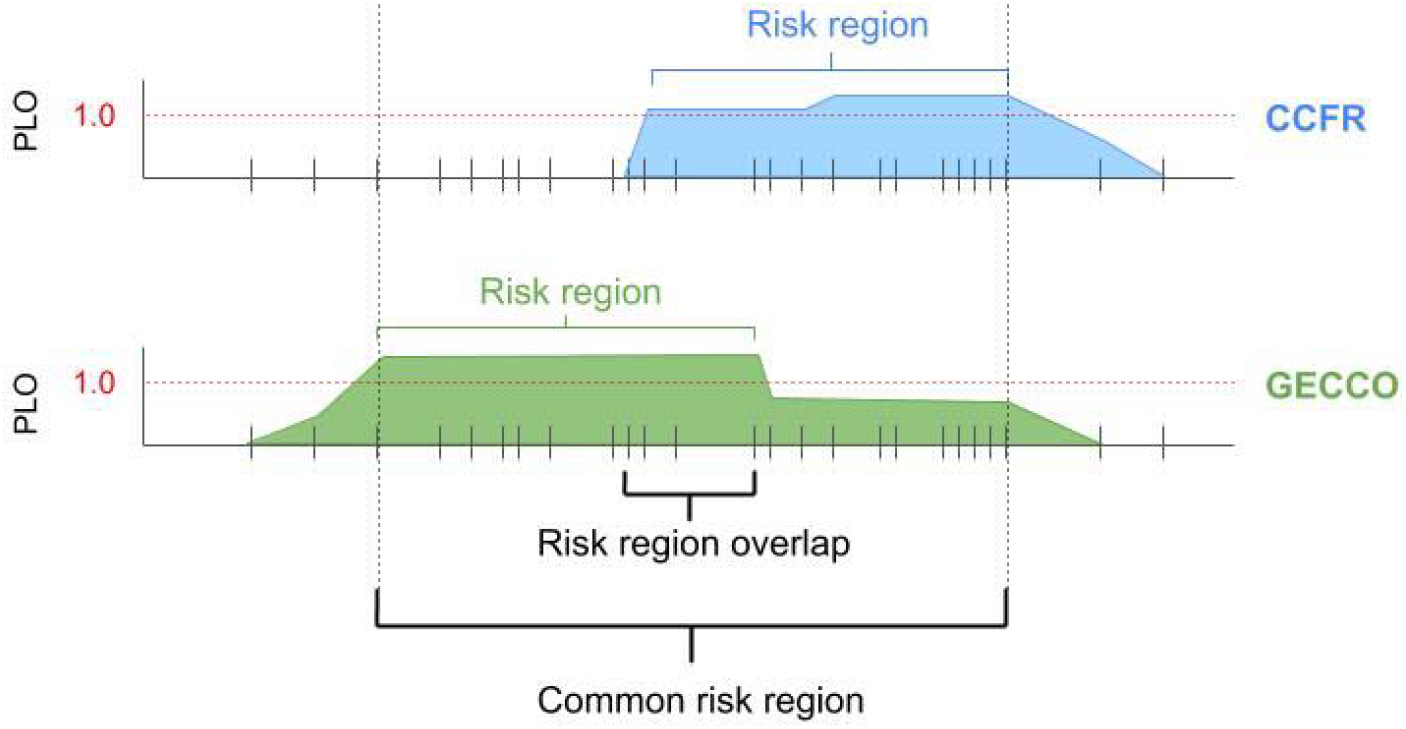
Risk regions were defined as having a posterior log-odds (PLO) above one (red horizontal dotted line). Common risk regions were defined as the boundary SNPs with a PLO above one in either CCFR and GECCO data sets (black vertical dotted line), and not the boundary SNPs with a PLO above one in both CCFR and GECCO data sets (risk region overlap). This approach was taken to ensure that risk regions are not necessarily restricted for DEPTH stage 2 and 3 analyses.

**Supplementary Figure 3.**
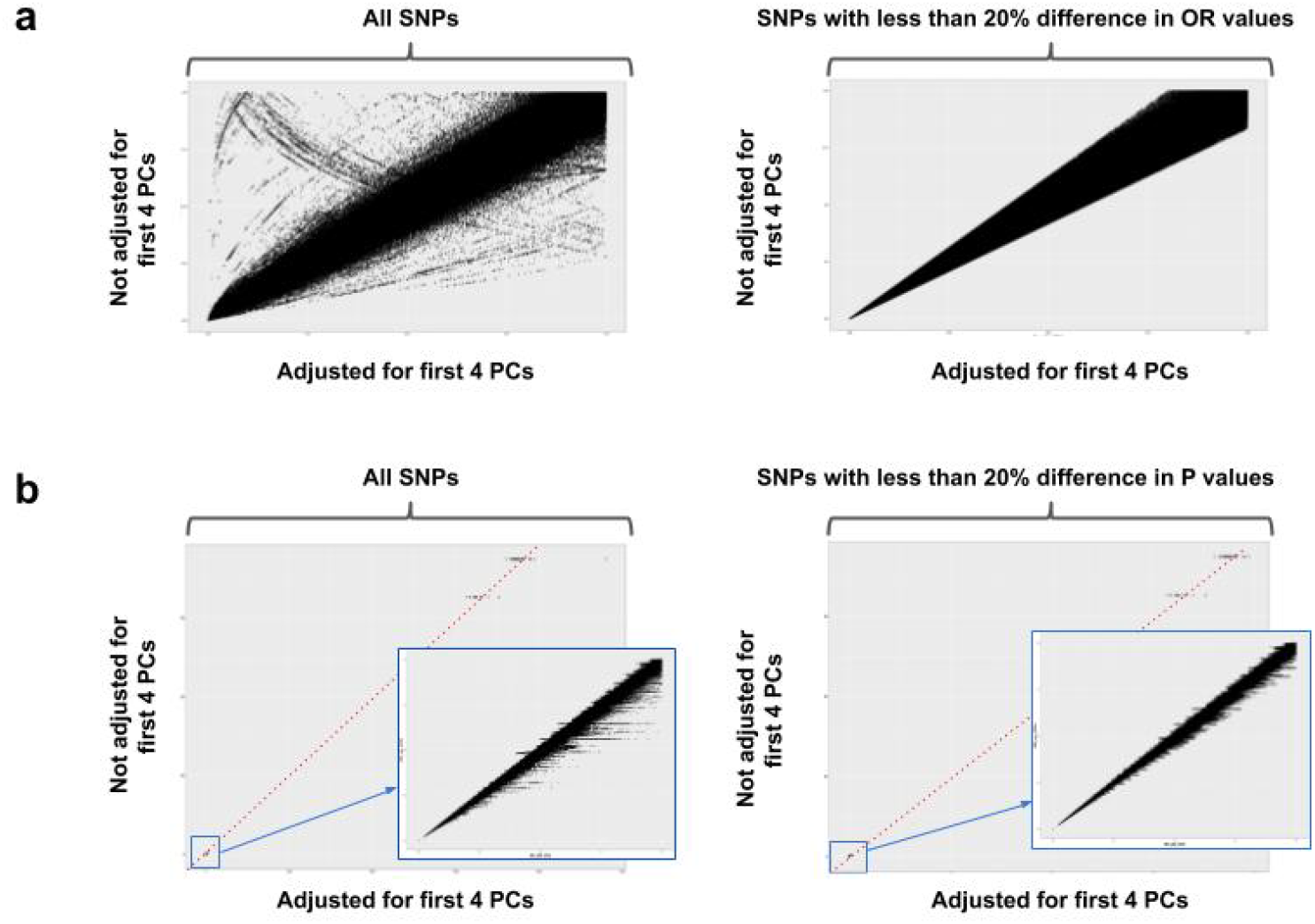
Following logistic regression analyses that adjusted for (*x*-axis), or did not adjust for (*y*-axis), the first four principal components (PCs), SNPs were retained if the odds ratios ***(a)*** or ***(b)*** P values did not differ by more than 20% (graphs on the right column).

**Supplementary Figure 4.**
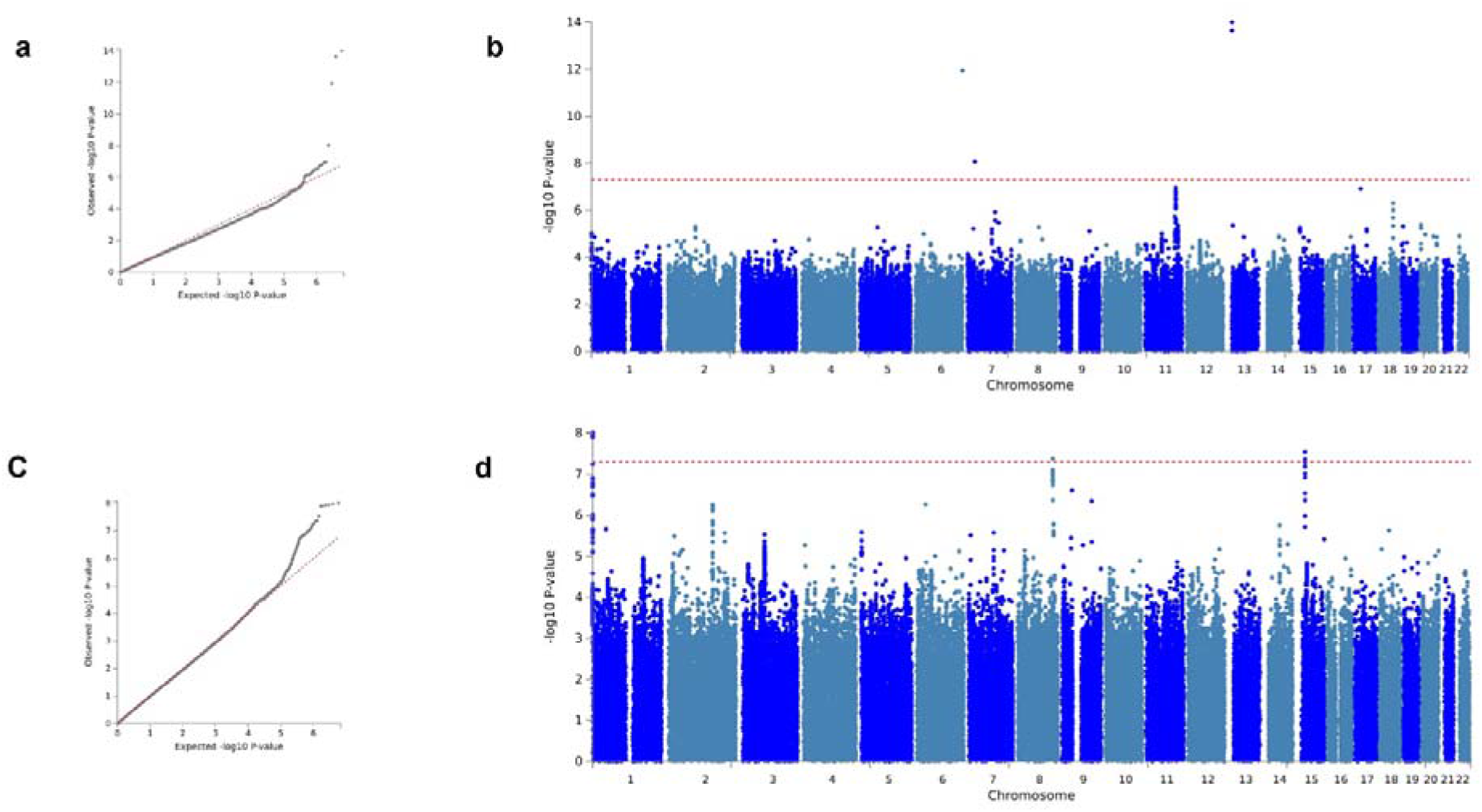
QQ plots of ***(a)*** CCFR and ***(b)*** GECCO from logistic regression analyses. Manhattan plots of ***(c)*** CCFR and ***(d)*** GECCO from logistic regression analyses, with the points above the dotted red line representing SNPs that are significant at *P* < 5×10^−8^.

## Author’s Contributions

AGRF is supported by the Australian Government National Collaborative Research Infrastructure Initiative through Bioplatforms Australia.

## Acknowledgements

The Colon Cancer Family Registry (CCFR, www.coloncfr.org) is supported in part by funding from the National Cancer Institute (NCI), National Institutes of Health (NIH) (award U01 CA167551). Support for case ascertainment was provided in part from the Surveillance, Epidemiology, and End Results (SEER) Program and the following U.S. state cancer registries: AZ, CO, MN, NC, NH; and by the Victoria Cancer Registry (Australia) and Ontario Cancer Registry (Canada). The CCFR Set-1 (Illumina 1 M/1 M-Duo) and Set-2 (Illumina Omni1-Quad) scans were supported by NIH awards U01 CA122839 and R01 CA143247 (to GC). The CCFR Set-3 (Affymetrix Axiom CORECT Set array) was supported by NIH award U19 CA148107 and R01 CA81488 (to SBG). The CCFR Set-4 (Illumina OncoArray 600 K SNP array) was supported by NIH award U19 CA148107 (to SBG) and by the Center for Inherited Disease Research (CIDR), which is funded by the NIH to the Johns Hopkins University, contract number HHSN268201200008I.

Genetics and Epidemiology of Colorectal Cancer Consortium (GECCO) is supported in part by funding from: National Cancer Institute, National Institutes of Health, U.S. Department of Health and Human Services (U01 CA164930, U01 CA137088, R01 CA059045). Genotyping/Sequencing services were provided by the Center for Inherited Disease Research (CIDR) (X01-HG008596 and X-01-HG007585). This research was funded in part through the NIH/NCI Cancer Center Support Grant P30 CA015704.

ASTERISK is supported in part by funding from: a Hospital Clinical Research Program (PHRC-BRD09/C) from the University Hospital Center of Nantes (CHU de Nantes) and supported by the Regional Council of Pays de la Loire, the Groupement des Entreprises Françaises dans la Lutte contre le Cancer (GEFLUC), the Association Anne de Bretagne Génétique and the Ligue Régionale Contre le Cancer (LRCC).

DACHS is supported in part by funding from the German Research Council (BR 1704/6-1, BR 1704/6-3, BR 1704/6-4, CH 117/1-1, HO 5117/2-1, HE 5998/2-1, KL 2354/3-1, RO 2270/8-1 and BR 1704/17-1), the Interdisciplinary Research Program of the National Center for Tumor Diseases (NCT), Germany, and the German Federal Ministry of Education and Research (01KH0404, 01ER0814, 01ER0815, 01ER1505A and 01ER1505B).

Harvard cohorts (HPFS, NHS, PHS) are supported in part by funding from: HPFS is supported by the National Institutes of Health (P01 CA055075, UM1 CA167552, U01 CA167552, R01 CA137178, R01 CA151993, R35 CA197735, K07 CA190673, and P50 CA127003), NHS by the National Institutes of Health (R01 CA137178, P01 CA087969, UM1 CA186107, R01 CA151993, R35 CA197735, K07CA190673, and P50 CA127003) and PHS by the National Institutes of Health (R01 CA042182).

MEC is supported in part by funding from: National Institutes of Health (R37 CA54281, P01 CA033619, and R01 CA063464).

OFCCR is supported in part by funding from: The Ontario Familial Colorectal Cancer Registry was supported in part by the National Cancer Institute (NCI) of the National Institutes of Health (NIH) under award U01 CA167551 and award U01/U24 CA074783 (to SG). Additional funding for the OFCCR and ARCTIC testing and genetic analysis was through and a Canadian Cancer Society CaRE (Cancer Risk Evaluation) program grant and Ontario Research Fund award GL201-043 (to BWZ), through the Canadian Institutes of Health Research award 112746 (to TJH), and through generous support from the Ontario Ministry of Research and Innovation.OSUMC: OCCPI funding was provided by Pelotonia and HNPCC funding was provided by the NCI (CA16058 and CA67941).

SCCFR is supported in part by funding from: The Seattle Colon Cancer Family Registry was supported in part by the National Cancer Institute (NCI) of the National Institutes of Health (NIH) under awards U01 CA167551, U01 CA074794 (to JDP), and awards U24 CA074794 and R01 CA076366 (to PAN).

VITAL is supported in part by funding from: National Institutes of Health (K05 CA154337). WHI is supported in part by funding from: the National Heart, Lung, and Blood Institute, National Institutes of Health, U.S. Department of Health and Human Services through contracts HHSN268201100046C, HHSN268201100001C, HHSN268201100002C, HHSN268201100003C, HHSN268201100004C, and HHSN271201100004C.

## Acknowledgements

ASTERISK: We are very grateful to Dr. Bruno Buecher without whom this project would not have existed. We also thank all those who agreed to participate in this study, including the patients and the healthy control persons, as well as all the physicians, technicians and students.

DACHS: We thank all participants and cooperating clinicians, and Ute Handte-Daub, Utz Benscheid, Muhabbet Celik and Ursula Eilber for excellent technical assistance.

Harvard cohorts (HPFS, NHS, PHS): The study protocol was approved by the institutional review boards of the Brigham and Women’s Hospital and Harvard T.H. Chan School of Public Health, and those of participating registries as required. We would like to thank the participants and staff of the HPFS, NHS and PHS for their valuable contributions as well as the following state cancer registries for their help: AL, AZ, AR, CA, CO, CT, DE, FL, GA, ID, IL, IN, IA, KY, LA, ME, MD, MA, MI, NE, NH, NJ, NY, NC, ND, OH, OK, OR, PA, RI, SC, TN, TX, VA, WA, WY. The authors assume full responsibility for analyses and interpretation of these data.

SCCFR: The authors would like to thank the study participants and staff of the Hormones and Colon Cancer and Seattle Cancer Family Registry studies (CORE Studies).

WHI: The authors thank the WHI investigators and staff for their dedication, and the study participants for making the program possible. A full listing of WHI investigators can be found at: http://www.whi.org/researchers/Documents%20%20Write%20a%20Paper/WHI%20Investigator%20Short%20List.pdf.

## References

1. Australian Institute of H, Welfare. Cancer in Australia: Actual incidence data from 1982 to 2013 and mortality data from 1982 to 2014 with projections to 2017. Asia Pac J Clin Oncol 2018;14(1):5–15 doi 10.1111/ajco.12761.

2. Siegel RL, Miller KD, Jemal A. Cancer statistics, 2019. CA Cancer J Clin 2019;69(1):7–34 doi 10.3322/caac.21551.

3. Hardcastle JD, Chamberlain JO, Robinson MH, Moss SM, Amar SS, Balfour TW, et al. Randomised controlled trial of faecal-occult-blood screening for colorectal cancer. Lancet 1996;348(9040):1472–7 doi 10.1016/S0140-6736(96)03386-7.

4. Kronborg O, Fenger C, Olsen J, Jorgensen OD, Sondergaard O. Randomised study of screening for colorectal cancer with faecal-occult-blood test. Lancet 1996;348(9040):1467–71 doi 10.1016/S0140-6736(96)03430-7.

5. Mandel JS, Bond JH, Church TR, Snover DC, Bradley GM, Schuman LM, et al. Reducing mortality from colorectal cancer by screening for fecal occult blood. Minnesota Colon Cancer Control Study. N Engl J Med 1993;328(19):1365–71 doi 10.1056/NEJM199305133281901.

6. Zauber AG, Winawer SJ, O’Brien MJ, Lansdorp-Vogelaar I, van Ballegooijen M, Hankey BF, et al. Colonoscopic polypectomy and long-term prevention of colorectal-cancer deaths. N Engl J Med 2012;366(8):687–96 doi 10.1056/NEJMoa1100370.

7. Keum N, Giovannucci E. Global burden of colorectal cancer: emerging trends, risk factors and prevention strategies. Nat Rev Gastroenterol Hepatol 2019 doi 10.1038/s41575-019-0189-8.

8. Orlando G, Law PJ, Palin K, Tuupanen S, Gylfe A, Hanninen UA, et al. Variation at 2q35 (PNKD and TMBIM1) influences colorectal cancer risk and identifies a pleiotropic effect with inflammatory bowel disease. Human molecular genetics 2016;25(11):2349–59 doi 10.1093/hmg/ddw087.

9. Tanikawa C, Kamatani Y, Takahashi A, Momozawa Y, Leveque K, Nagayama S, et al. GWAS identifies two novel colorectal cancer loci at 16q24.1 and 20q13.12. Carcinogenesis 2018;39(5):652–60 doi 10.1093/carcin/bgy026.

10. Law PJ, Timofeeva M, Fernandez-Rozadilla C, Broderick P, Studd J, Fernandez-Tajes J, et al. Association analyses identify 31 new risk loci for colorectal cancer susceptibility. Nat Commun 2019;10(1):2154 doi 10.1038/s41467-019-09775-w.

11. Huyghe JR, Bien SA, Harrison TA, Kang HM, Chen S, Schmit SL, et al. Discovery of common and rare genetic risk variants for colorectal cancer. Nature genetics 2019;51(1):76–87 doi 10.1038/s41588-018-0286-6.

12. Schmit SL, Edlund CK, Schumacher FR, Gong J, Harrison TA, Huyghe JR, et al. Novel Common Genetic Susceptibility Loci for Colorectal Cancer. Journal of the National Cancer Institute 2019;111(2):146–57 doi 10.1093/jnci/djy099.

13. Makalic E, Schmidt DF, Hopper JL. DEPTH: A Novel Algorithm for Feature Ranking with Application to Genome-Wide Association Studies. AI 2013: Advances in Artificial Intelligence; 2013; Cham. Springer International Publishing. p 80-5. (AI 2013: Advances in Artificial Intelligence).

14. MacInnis RJ, Schmidt DF, Makalic E, Severi G, FitzGerald LM, Reumann M, et al. Use of a Novel Nonparametric Version of DEPTH to Identify Genomic Regions Associated with Prostate Cancer Risk. Cancer epidemiology, biomarkers & prevention : a publication of the American Association for Cancer Research, cosponsored by the American Society of Preventive Oncology 2016;25(12):1619–24 doi 10.1158/1055-9965.EPI-16-0301.

15. Schumacher FR, Schmit SL, Jiao S, Edlund CK, Wang H, Zhang B, et al. Genome-wide association study of colorectal cancer identifies six new susceptibility loci. Nat Commun 2015;6:7138 doi 10.1038/ncomms8138.

16. Peters U, Jiao S, Schumacher FR, Hutter CM, Aragaki AK, Baron JA, et al. Identification of Genetic Susceptibility Loci for Colorectal Tumors in a Genome-Wide Meta-analysis. Gastroenterology 2013;144(4):799–807 e24 doi 10.1053/j.gastro.2012.12.020.

17. Purcell S, Neale B, Todd-Brown K, Thomas L, Ferreira MA, Bender D, et al. PLINK: a tool set for whole-genome association and population-based linkage analyses. Am J Hum Genet 2007;81(3):559–75 doi 10.1086/519795.

18. Lafayette L, Sauter G, Vu L, Meade B. Spartan Performance and Flexibility: An HPC-Cloud Chimera. OpenStack Summit. Barcelona 2016.

19. Goldman M, Craft B, Swatloski T, Ellrott K, Cline M, Diekhans M, et al. The UCSC Cancer Genomics Browser: update 2013. Nucleic Acids Res 2013;41(Database issue):D949–54 doi 10.1093/nar/gks1008.

20. Watanabe K, Taskesen E, van Bochoven A, Posthuma D. Functional mapping and annotation of genetic associations with FUMA. Nat Commun 2017;8(1):1826 doi 10.1038/s41467-017-01261-5.

21. Risch N, Merikangas K. The future of genetic studies of complex human diseases. Science 1996;273(5281):1516–7 doi 10.1126/science.273.5281.1516.

22. Shiina T, Hosomichi K, Inoko H, Kulski JK. The HLA genomic loci map: expression, interaction, diversity and disease. J Hum Genet 2009;54(1):15–39 doi 10.1038/jhg.2008.5.

23. Christofi T, Baritaki S, Falzone L, Libra M, Zaravinos A. Current Perspectives in Cancer Immunotherapy. Cancers (Basel) 2019;11(10) doi 10.3390/cancers11101472.

